# Evidence on WASH interventions in Negelle-Arsi District, Oromia Regional State, Ethiopia: a cross-sectional data analysis

**DOI:** 10.64898/2026.04.08.26349166

**Authors:** Walelign Fetahei, Birhanu Yadete

## Abstract

**BACKGROUND:** Integrated WASH interventions play a vital role in addressing public health challenges in communities. Governments and development partners work collaboratively to improve access to safe water, sanitation, and hygiene services in order to mitigate these challenges. This study assesses the impact of WASH intervention on water access and supply, as well as changes in participants’ knowledge, attitudes, and practices in rural communities, using cross-sectional data by comparing treatment and control groups.

**METHODS:** The study used cross-sectional data collected in May 2025 from six KAs, three intervention and three non-intervention areas, through a structured household questionnaire. A total of 396 households with children under five were included in the survey, equally divided between the treatment and control groups. Descriptive analysis was used to assess the differences among the groups in water access and supply as well as to examine the differences in knowledge, attitudes, and practices of households participating in the study.

**RESULTS:** The prevalence of diarrhea among children under five years old in the rural setting during the two weeks prior to data collection showed that approximately 69 children (34.9 percent) in the control group and 5 children in the treated group (2.5 percent) experienced diarrhea. Treated households had better child health: more facility births (88.9% vs. 63.6%), higher breastfeeding (98% vs. 89.9%) and vaccination coverage (78.8% vs. 59.1%), and lower diarrhea incidence (8.1% vs. 37.4%). Water access was improved, with all treated households using protected sources, shorter collection times (13.85 vs. 55.84 minutes), and higher consumption (20.6 vs. 10.5 liters/day). Safely managed water services were 59.6% in treated vs. 1% in control households. Sanitation and hygiene also improved: latrine access (95% vs. 78.3%), open defecation (23.2% vs. 52%), and handwashing with soap (48.5% vs. 12.1%). Knowledge, attitudes, and practices were higher in treated households: good WASH knowledge (91.4% vs. 70%), favorable attitudes (99% vs. 74.8%), and proper WASH practices (85.4% vs. 26.3%).

**CONCLUSION:** The findings confirm that integrated WASH interventions, combining improved water infrastructure with sanitation promotion, hygiene education, and community engagement, can substantially improve household WASH conditions and reduce waterborne diseases. Sustaining these gains will require continued investment in water infrastructure, strengthening community awareness, and expanding behavior change initiatives to other communities.

## Background

The concept of Water, Sanitation, and Hygiene (WASH) is a central idea for public health and human development to ensure a healthy environment and well-being of human existence. It comprises access to safe drinking water, proper sanitation facilities, and hygiene practices that WASH interventions aim to strive to prevent disease transmission and ultimately improve health outcomes of the communities.

Water is the most indispensable natural resource in the world for every living being. The entire life-support systems depend on this vital resource. It is crucial to all living entities as it is directly consumed by them [22]. The word hygiene mainly focuses on diseases and health, while sanitation focuses on the safe disposal of human waste, which could be human urine and feces. However, both hygiene and sanitation aim at creating a disease-free world that is full of healthy people [13].

Morbidity is the state of being diseased or unhealthy within a population, encompassing the incidence, prevalence, and severity of illnesses or health conditions [17]. Diarrhea morbidity refers to the occurrence or prevalence of diarrheal within a population, indicating the burden of illness caused by frequent loose or watery stools, often resulting from infections transmitted through contaminated water, poor sanitation, or inadequate hygiene practices [25].

Ethiopia lags behind least developing countries (LDCs) in drinking water supply coverage, sanitation, and hygiene practices. Specifically, the water coverage is promising, with 79% of the population having basic water supply compared to 81% for LDCs in 2022. However, in sanitation and hygiene practices, it is extremely below fourfold that of the LDCs. The sanitation and hygiene coverage of LDCs are 42% and 34%, respectively, whereas that of Ethiopia is 9% and 8% in 2022, respectively [21].

In 2022, over 74% of the total population (about 73 million people) had access to drinking water from improved sources in Ethiopia. Access to improved water is higher in urban areas than in rural areas. However, access in rural areas increased by seven percentage points from 62% in 2019 to 69% in 2022, narrowing the rural-urban gap [11].

Similarly, in 2022, only 26% of the population (about 23.5 million people) had improved sanitation facilities in Ethiopia. The majority, 74% of the population (about 74.8 million people), relied on unimproved sanitation facilities or open defecation. Access to improved sanitation facilities was 58% in urban areas and 15% in rural areas. Regarding hygiene, only one-third of households had a handwashing facility in their residence. Moreover, the prevalence of basic hygiene services is very low in the country and worse in rural areas (4%). The availability of a handwashing facility is more common in urban areas (55%) than in rural areas (22%0 [11].

Worldwide, unsafe water, inadequate sanitation, or insufficient hygiene leads to 80% of diarrhoea cases [16]. A significant proportion of diarrhea can be prevented through safe drinking water and adequate sanitation and hygiene [25, 26].

Recent evidence shows that inadequate water, sanitation, and hygiene (WASH) services remain a major contributor to the global disease burden, particularly among young children [19].

Multiple studies across different low-resource settings consistently show that poor water, sanitation, and hygiene (WASH) knowledge and practices are strongly linked to higher childhood morbidity and undernutrition. Handwashing promotion alone has been shown to reduce diarrhea incidence, whereas broader WASH interventions often show limited impact [12]. In rural Bangladesh, Ethiopia, Nepal, India, and Niger, children under five experienced higher rates of diarrhea and other illnesses when caregivers had low WASH knowledge, lacked sanitation facilities, or practiced unsafe hygiene, with education level, access to latrines, handwashing habits, and safe water use emerging as key determinants [2, 3, 4, 5, 7, 14, 15, 16, 18, 20]. Factors such as improper waste disposal, lack of vaccination, unsafe complementary feeding, and inadequate water storage further exacerbated diarrheal risk, highlighting that targeted education, improved hygiene practices, and basic sanitation are critical for reducing childhood disease and improving nutritional outcomes.

## Methods

### Design and setting of the study

A cross-sectional study based on a WASH project intervention was conducted in Negelle-Arsi District, Oromia Regional State, Ethiopia. Descriptive analysis was employed to examine the relationships between variables among the intervention (treated) and non-intervention (control) groups.

The project employed integrated water supply, sanitation and hygiene (WASH) intervention coupled with community-led total sanitation and hygiene (CLTSH) approach to bring health outcomes in the community. These programs ensure that water supply systems, sanitation facilities, and hygiene education are all integrated and managed together to maximize health benefits (Andrés et al, 2018). Interventions to improve access to clean water, sanitation facilities, and hygiene behaviors (WASH) represent key opportunities to improve child health and well-being by preventing the spread of infectious diseases and improving nutritional status [9].

### Data sources and types

The study collected quantitative primary data from rural households in the selected intervention and non-intervention Kebele Administrations (KAs) using a structured survey questionnaire. Before the main data collection, the questionnaire was pre-tested in the field to ensure its clarity and reliability. Data collectors with prior survey experience were recruited and received comprehensive training to familiarize themselves with the questionnaire and the overall data collection process.

### Sampling design and techniques

The study employed a probability-based random sampling technique to select participants from the sampling frame. In general, a two-stage random sampling procedure was used to identify and select the respondent households included in the study.

In the first stage, three Kebele Administrations (KAs) were randomly selected from both intervention and non-intervention areas, making a total of six KAs. Namely: ***Kersa Elala, Gorbi Derera, and Danshe*** were selected from the intervention areas, while ***Hada Bila, Hada Bosso, and Chri Lalu*** were selected from the non-intervention areas using the lottery method. In the second stage, households were selected using systematic sampling with a fixed interval (n). The first household was chosen randomly from the initial interval, and subsequent households were selected by adding the sampling interval (n). Finally, the number of households sampled from each KA was determined proportionally, based on the size of the population in each cluster kebele.

**Figure 1:**
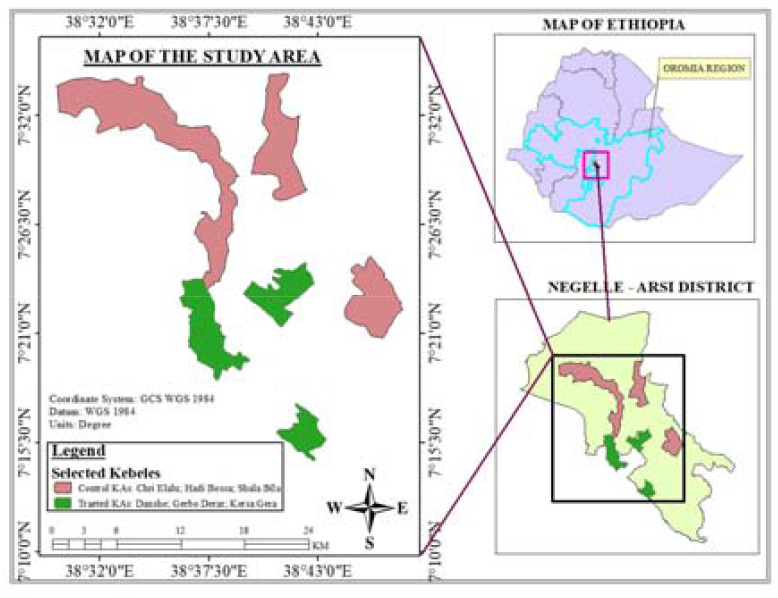
Map of the study area.

### Sample size determination

The study targeted households that participated in the WASH intervention program, along with a control group of comparable households from non-beneficiary kebele administrations. The study employed the Yemane formula to determine the sample size, ensuring a representative subset of the population without requiring a full census [27]. The sample size was determined using the Yemane formula, as presented below:

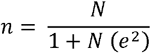

Where “n” is the sample size, “N” is the total population, and “e” is the level of precision or margin of error. Using the above formula, the sample size was calculated from six kebele administrations (three intervention and three non-intervention) based on the identified sampling frame. The total population (N) was 44,156, with a precision level (e) of 5%. Accordingly, a total of 396 households were selected, comprising 198 households in the intervention group and 198 in the control group 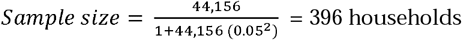 This sample size was allocated to each kebele administration using probability proportional to the size of their respective sampling frames, as presented in Table 1.

**Table 1:** Study Sample Size and Sample Frame.

|  | Sampled KAs | Population |  | Sample Size |  |
| --- | --- | --- | --- | --- | --- |
|  |  | Treated | Control | Treated | Control |
| Treated KAs | Kersa Elala | 6,736 |  | 60 |  |
|  | Gorbi Derara | 8,409 |  | 71 |  |
|  | Danshe | 7,861 |  | 67 |  |
| Control KAs | Shala Bila |  | 6,661 |  | 62 |
|  | Hada Bosso |  | 5,922 |  | 56 |
|  | Chri Lalu |  | 8,567 |  | 80 |
| **Sub-Total** |  | **23,006** | **21,150** | **198** | **198** |
| **Grand Total** |  | **44,156** |  | **396** |  |

Source: Negelle-Arsi District Health Office, 2024

### Operational Definitions and Measurements

#### Safe Drinking Water

It is defined as water from an improved water source, which includes household connections, public standpipes, boreholes, protected dug wells, protected springs and rainwater collections. Similarly, access to safe drinking water is defined as the availability of at least 20 liters per person per day from an improved source within 1 kilometer of the user’s dwelling [21].

***Sanitation*** is defined as access to and use of facilities and services for the safe disposal of human excreta [23].

***Hygiene*** is the conditions and practices that help maintain health and prevent the spread of disease including handwashing, food hygiene, and menstrual hygiene management [23].

***Diarrheal Disease*** is defined as the passage of three or more loose or liquid stools per day, or more frequent passage than is normal for the individual [24].

#### Knowledge Level

Respondents who obtained a mean knowledge score of ≤ 0.50 were categorized as having poor knowledge, whereas those scoring > 0.50 were categorized as having good knowledge.

#### Attitude Level

A mean attitude score of ≤ 0.50 indicated a negative attitude, while a score above 0.50 indicated a positive attitude.

#### Practice Level

Participants with a mean practice score of ≤ 0.50 were considered to have poor practice, and those with scores > 0.50 were considered to have good practice.

#### Data analysis

Data were collected from respondents using the Kobo Toolbox platform. The collected data were subsequently cleaned and prepared in Stata 17 by running tabulations for each variable to identify and address missing values, inconsistencies, outliers, and data entry errors. A descriptive analysis was conducted to examine the socio-demographic, socio-economic, and maternal characteristics of both the treatment and control groups. The water supply, sanitation, and hygiene status of the participants were also assessed. In addition, respondents’ knowledge, attitudes, and practices related to WASH were analyzed using descriptive statistics

#### Quality Assurance

All data collectors had a bachelor’s degree and prior experience in similar data collection activities. The questionnaire was pre-tested with 20 respondents to ensure it captured the necessary information and verified its reliability. The pre-test results confirmed the instrument’s suitability for the study. Data quality was further enhanced using the Kobo Toolbox platform, which included built-in quality control features. GPS coordinates were recorded at each respondent’s household, and photos of the house or latrine or handwashing facility were taken with consent when appropriate. Data were submitted daily through the online system and regularly checked, especially the coordinates, to confirm the respondents’ actual location and data quality.

#### Ethical Considerations

The study was conducted in line with the principles of the 1964 Helsinki Declaration and its 2024 revision, as well as other comparable ethical standards [6]. Prior to the data collection, official permission to carry out the study was obtained from the local government through written authorization. The data collectors maintained confidentiality when gathering information from participants. While the respondents were identifiable for the study, their privacy was protected through an anonymous approach. The study followed ethical guidelines by obtaining informed consent, ensuring confidentiality, preventing harm and conflicts of interest, and respecting the local community’s beliefs and customs. The research also complied with local regulations and governance frameworks.

## Results

### Household characteristics of respondents

The study included 396 households from six Kebele administrations. Among the respondents, 9% (34 households) were female-headed, while the majority, 91% (362 households), were male-headed. Female-headed households were more common in the control group (25 households) than in the treated group (9 households).

As shown in Table 2, the two groups exhibit statistically significant differences in terms of family size, sex, and education level of the household head. The average family size of the control group, 6.47, is higher than that of the treated group, 5.69. Regarding the education level of the household head, 25.3 percent of the control group did not read and write, in contrast with the treated group at 7.5 percent. Additionally, a significant number of the treated group, 62.7 percent, attended formal education (primary to tertiary) compared to the control group at 45 percent.

**Table 2:**
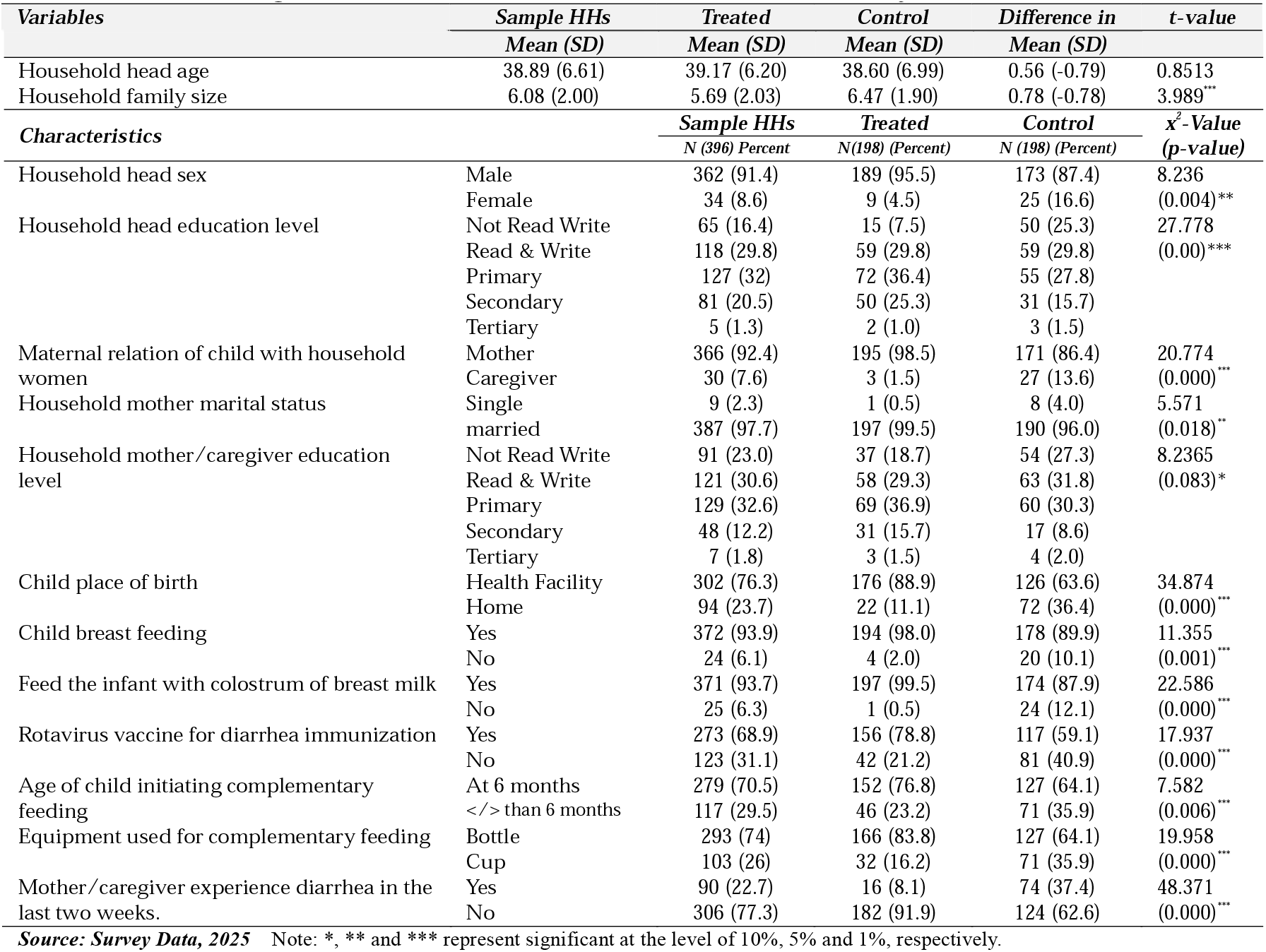
Background Characteristics of households and under-five year children (n=396)

Almost all mothers /caregivers in the treated group were married (99.5%), compared to 96% in the control group. Illiteracy was higher in the control group (27.3%) than in the treated group (18.7%). Likewise, a larger share of respondents in the treated group (54.1%) had attended formal education, compared to 40% in the control group.

### Under-Five Children Characteristics

As shown in Table 2, there are statistically significant differences between the treated and control groups across all under-five child variables. A larger share of mothers in the treated group (88.9%) gave birth in a health facility compared to those in the control group (63.6%). Breastfeeding (98%) and colostrum feeding (99.5%) were also more common in the treated group than in the control group (89.9% and 87.9%, respectively).

Similarly, rotavirus vaccination coverage was higher among children in the treated group (78.8%) than in the control group (59.1%). Complementary feeding at the recommended age of six months was reported by 76.8% of respondents in the treated group and 64.1% in the control group. Bottle feeding was more common in the treated group (83.8%), while cup use was higher in the control group (35.9%).

The incidence of diarrhea was notably higher in the control group, with 37.4% of children experiencing diarrhea in the two weeks prior to the survey, compared to 8.1% in the treated group.

### Household Water Supply and Access

All water access and supply variables are statistically significant at the 1% level, indicating clear differences between the treated and control groups due to the water supply intervention as shown in Table 7. All respondents in the treated group had access to protected water sources, compared to 76.8% in the control group. Similarly, 90.4% of the treated group reported that their water sources were reliable and sufficient for their needs, while only 60.6% of the control group said the same, with 39.4% reporting unreliable sources.

Average daily water consumption was 20.6 liters per person in the treated group, compared to 10.51 liters in the control group. The average round-trip time to fetch water, including queuing, was 13.85 minutes for the treated group and 55.84 minutes for the control group. This means the treated group met the international standard of collecting water within 30 minutes, while the control group spent significantly more time. In addition, households in the treated group stored drinking water for an average of 1.73 days, compared to 2.22 days in the control group.

As indicated in Table 3 above, a large majority of respondents in the treated group (96%) stored drinking water separately, compared to 61.2% in the control group. Most households in both groups used jerrycan containers for water storage (96% in the treated group and 72% in the control group), followed by buckets and pots /jars. Water was mainly accessed by pouring rather than dipping, with only a small proportion using dipping (4.8% in the treated group and 0.5% in the control group).

**Table 3:** Water Access and Supply Variables (n=396)

| Variables |  | Sample HHs | Treated | Control | Difference in | t-value |
| --- | --- | --- | --- | --- | --- | --- |
|  |  | Mean (SD) | Mean (SD) | Mean (SD) | Mean (SD) |  |
| Water consumption in liter per person per day |  | 15.34 (7.65) | 20.60 (6.09) | 10.08 (4.63) | 10.51 (1.46) | 18.80*** |
| Time taken to fetch H<sub>2</sub>O in minute |  | 34.85 (38.73) | 13.85 (10.33) | 55.84 (44.88) | -41.99 (-34.55) | 12.83*** |
| Water storage in house in days |  | 1.97 (1.01) | 1.73 (0.66) | 2.22 (1.22) | -0.49 (-0.56) | -4.94*** |
| Characteristics |  | Sample HHs | Treated | Control | x<sup>2</sup>-Value (p-value) |  |
|  |  | N (396) Percent | N(198) (Percent) | N (198) (Percent) |  |  |
| Water source for the last one year | Protected | 350 (88.4) | 198 (100) | 152 (76.8) | 52.045 |  |
|  | Unprotected | 46 (11.6) | 0 (0) | 46 (23.2) | (0.000)*** |  |
| Water source reliability in terms of availability | Yes | 299 (75.5) | 179 (90.4) | 120 (60.6) | 47.529 |  |
|  | No | 97 (24.5) | 19 (9.6) | 78 (39.4) | (0.000)*** |  |
| Store drinking water separately | Yes | 323 (81.6) | 190 (96) | 133 (61.2) | 54.566 |  |
|  | No | 173 (18.4) | 8 (4) | 65 (32.8) | (0.000)*** |  |
| Pouring water in the house for use | By Pouring | 377 (95.2) | 197 (99.5) | 180 (90.9) | 15.977 |  |
|  | By Dipping | 19 (4.8) | 1 (0.5) | 18 (9.1) | (0.000)*** |  |
| Practice of water treatment before use | Yes | 15 (3.8) | 2 (1) | 13 (6.6) | 8.384 |  |
|  | No | 381 (96.2) | 196 (99) | 185 (93.4) | (0.004)** |  |
| Level of drinking water services based on JMP ladder | Safely managed | 120 (30.3) | 118 (59.6) | 2 (1) | 188.234 |  |
|  | Basic | 107 (27.0) | 26 (13.1) | 81 (40.9) | (0.000)*** |  |
|  | Limited | 123 (31.1) | 54 (27.3) | 69 (33.9) |  |  |
|  | Unimproved | 40 (10.1) | 0 (0) | 40 (20.2) |  |  |
|  | Surface water | 6 (1.5) | 0 (0) | 6 (3.0) |  |  |

Source: Survey Data, 2025 Note: \*, \*\* and \*\*\* represent significant at the level of 10%, 5% and 1%, respectively.

Almost all respondents in the treated group (99%) did not treat water before use, mainly because they believed the water was safe. Similarly, 93.4% of the control group did not practice water treatment due to reasons such as lack of awareness, limited access to treatment options, or the perception that the water was safe. Overall, only 15 households (2 in the treated group and 13 in the control group) reported treating water using methods such as chlorine / Water Guard /Aqua Tabs, bleach, boiling, or filtration.

Regarding the level of drinking water services based on the UNICEF /WHO JMP ladder, 59.6 percent, 13.1 percent, and 27.3 percent of treated respondents have safely managed, basic, and limited levels of water services, respectively, which represent a significant level of access. In contrast, 1 percent, 40.9 percent, and 33.9 percent of control group respondents have safely managed, basic, and limited levels of water services, respectively. There is no unimproved or surface water access for the treated group, while 20.2 percent and 3 percent of control group members have unimproved and surface water experiences at the time of the study. This difference is a result of the intervention implemented in the treated community.

### Sanitation Situation of households

As shown in table 4, latrine access was higher in the treated group (95%) than in the control group (78.3%), with inaccessibility due to factors such as demolished or full latrines and inability to construct new ones. Open defecation was reported in 23.2% of treated households and 52% of control households, showing the intervention’s impact in reducing open defecation. Among households with latrines, 68.6% of the treated group and 39.4% of the control group had water and handwashing facilities at the toilet.

**Table 4:**
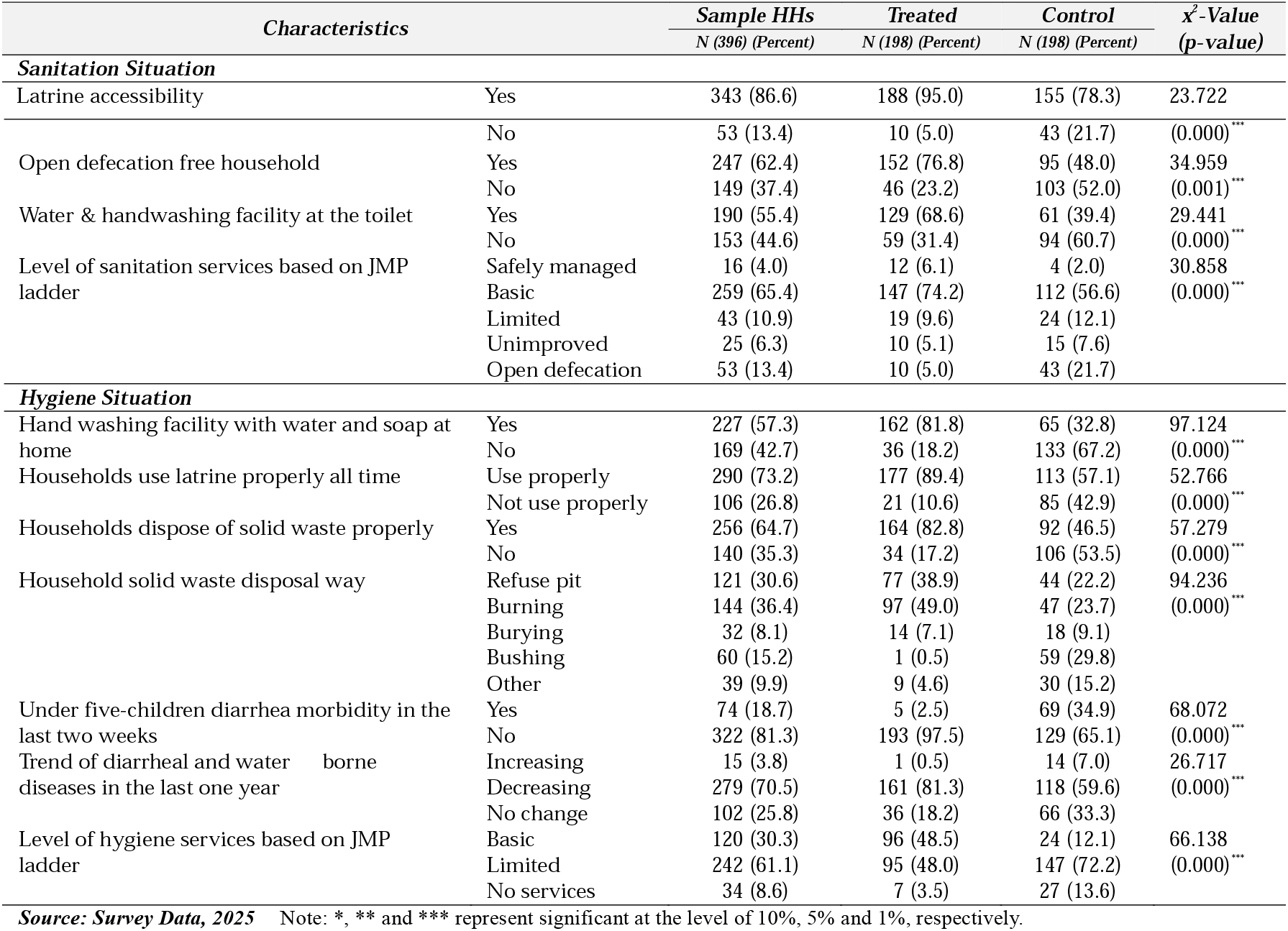
Sanitation and Hygiene Situation of households (n=396)

| **Characteristics** |  | **Sample HHs**<br>**N (396) (Percent)** | **Treated**<br>**N (198) (Percent)** | **Control**<br>**N (198) (Percent)** | **$\chi^2$-Value**<br>**(p-value)** |
| --- | --- | --- | --- | --- | --- |
| **Sanitation Situation** |  |  |  |  |  |
| Latrine accessibility | Yes | 343 (86.6) | 188 (95.0) | 155 (78.3) | 23.722 |
|  | No | 53 (13.4) | 10 (5.0) | 43 (21.7) | (0.000)*** |
| Open defecation free household | Yes | 247 (62.4) | 152 (76.8) | 95 (48.0) | 34.959 |
|  | No | 149 (37.4) | 46 (23.2) | 103 (52.0) | (0.001)*** |
| Water & handwashing facility at the toilet | Yes | 190 (55.4) | 129 (68.6) | 61 (39.4) | 29.441 |
|  | No | 153 (44.6) | 59 (31.4) | 94 (60.7) | (0.000)*** |
| Level of sanitation services based on JMP ladder | Safely managed | 16 (4.0) | 12 (6.1) | 4 (2.0) | 30.858 |
|  | Basic | 259 (65.4) | 147 (74.2) | 112 (56.6) | (0.000)*** |
|  | Limited | 43 (10.9) | 19 (9.6) | 24 (12.1) |  |
|  | Unimproved | 25 (6.3) | 10 (5.1) | 15 (7.6) |  |
|  | Open defecation | 53 (13.4) | 10 (5.0) | 43 (21.7) |  |
| **Hygiene Situation** |  |  |  |  |  |
| Hand washing facility with water and soap at home | Yes | 227 (57.3) | 162 (81.8) | 65 (32.8) | 97.124 |
|  | No | 169 (42.7) | 36 (18.2) | 133 (67.2) | (0.000)*** |
| Households use latrine properly all time | Use properly | 290 (73.2) | 177 (89.4) | 113 (57.1) | 52.766 |
|  | Not use properly | 106 (26.8) | 21 (10.6) | 85 (42.9) | (0.000)*** |
| Households dispose of solid waste properly | Yes | 256 (64.7) | 164 (82.8) | 92 (46.5) | 57.279 |
|  | No | 140 (35.3) | 34 (17.2) | 106 (53.5) | (0.000)*** |
| Household solid waste disposal way | Refuse pit | 121 (30.6) | 77 (38.9) | 44 (22.2) | 94.236 |
|  | Burning | 144 (36.4) | 97 (49.0) | 47 (23.7) | (0.000)*** |
|  | Burying | 32 (8.1) | 14 (7.1) | 18 (9.1) |  |
|  | Bushing | 60 (15.2) | 1 (0.5) | 59 (29.8) |  |
|  | Other | 39 (9.9) | 9 (4.6) | 30 (15.2) |  |
| Under five-children diarrhea morbidity in the last two weeks | Yes | 74 (18.7) | 5 (2.5) | 69 (34.9) | 68.072 |
|  | No | 322 (81.3) | 193 (97.5) | 129 (65.1) | (0.000)*** |
| Trend of diarrheal and water borne diseases in the last one year | Increasing | 15 (3.8) | 1 (0.5) | 14 (7.0) | 26.717 |
|  | Decreasing | 279 (70.5) | 161 (81.3) | 118 (59.6) | (0.000)*** |
|  | No change | 102 (25.8) | 36 (18.2) | 66 (33.3) |  |
| Level of hygiene services based on JMP ladder | Basic | 120 (30.3) | 96 (48.5) | 24 (12.1) | 66.138 |
|  | Limited | 242 (61.1) | 95 (48.0) | 147 (72.2) | (0.000)*** |
|  | No services | 34 (8.6) | 7 (3.5) | 27 (13.6) |  |

Source: Survey Data, 2025 Note: \*, \*\* and \*\*\* represent significant at the level of 10%, 5% and 1%, respectively.

Based on the UNICEF /WHO JMP ladder, most treated respondents had basic or higher sanitation services: 6.1% safely managed, 74.2% basic, and 9.6% limited. In the control group, 2% had safely managed, 56.6% basic, and 12.1% limited services. Unimproved and open defecation rates were higher in the control group (7.6% and 21.7%) than in the treated group (5.1% and 5.0%), reflecting the impact of the intervention.

### Hygiene Situation of households

Hygiene variables were significantly different between the groups at the 1% and 5% levels. About 48.5% of treated households had handwashing facilities with water and soap, compared to 12.1% in the control group. Proper latrine use was reported by 82.8% of treated respondents versus 55.1% of control respondents. Similarly, proper domestic waste disposal was higher in the treated group (82.8%) than in the control group (46.5%), with only 38.9% of treated and 22.2% of control respondents using refuse pits, while 29.8% of control households burned their waste, as displayed in Table 4.

Diarrhea incidence in the two weeks before the survey was 2.5% among children in the treated group and 34.9% in the control group. Regarding trends over the past year, 81.3% of treated respondents and 59.6% of control respondents reported a decrease in diarrhea and waterborne diseases, while a small share noted an increase (1% treated, 7% control).

Based on the UNICEF /WHO hygiene ladder, 48.5% of treated households and 12.1% of control households had basic hygiene (handwashing facility with soap and water). Limited access was reported in 48% of treated and 72% of control households (facility without soap or water), while 3.5% of treated and 13.6% of control households had no handwashing facility, representing a no-service level.

### Knowledge of households on WASH

Respondents’ WASH knowledge was assessed through several variables as indicated in Table 5 above. Most showed significant differences between the groups, except for awareness that water can be contaminated by liquid waste and its consequences. In the treated group, 89.9% recognized that unsafe water causes diarrhea and 91.4% understood the importance of using clean water for handwashing, compared to 74.2% and 60.1% in the control group. Similarly, 87.9% of treated respondents knew how animal dung can cause diseases, versus 65.7% in the control group. Knowledge of latrine necessity was higher in the treated group (94.4%) than in the control group (70.7%), and 92.4% of treated households understood the health risks of improper waste disposal, compared to 75.3% in the control group. Awareness of handwashing consequences was 74.8% in the treated group and 64.1% in the control group. Overall, 91.4% of treated households had good WASH knowledge, while about 70% of control households did, reflecting the impact of the project’s WASH campaign.

**Table 5:**
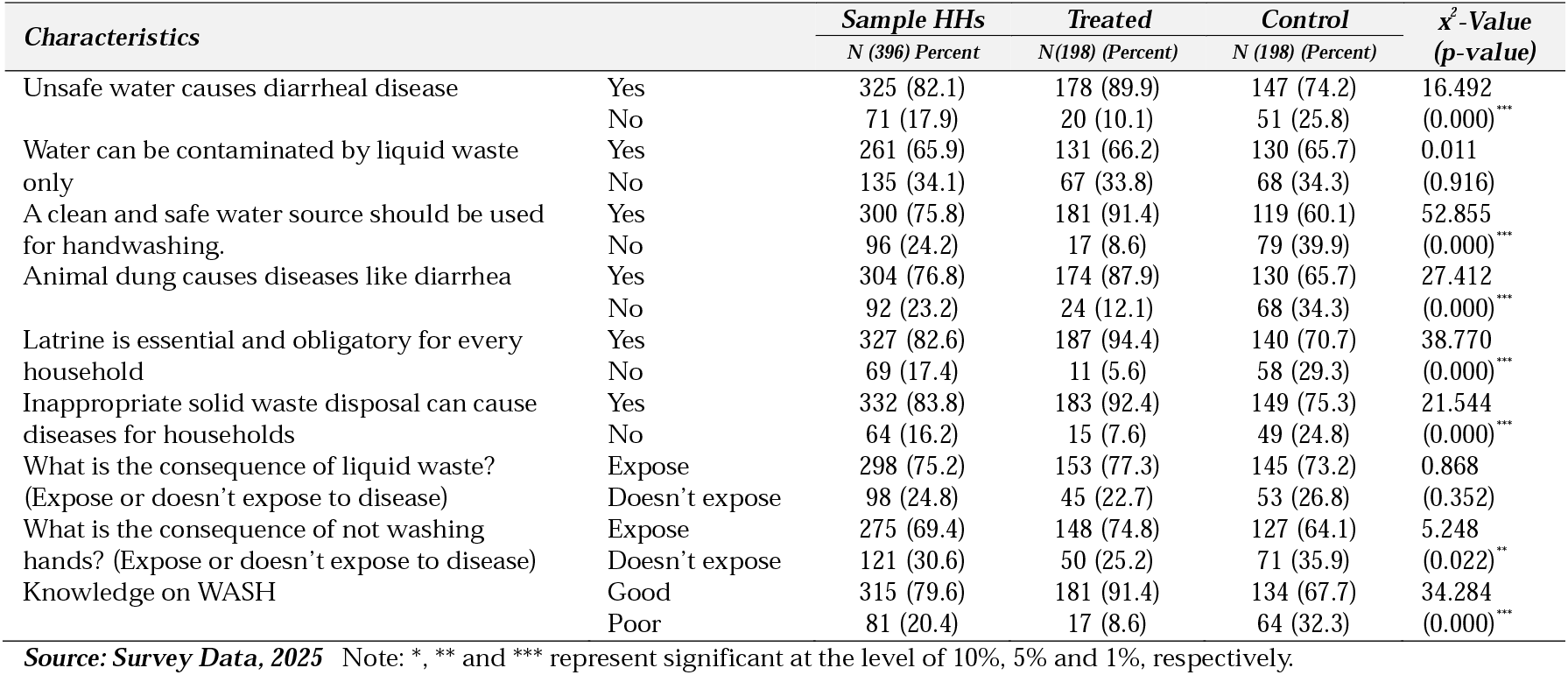
Knowledge of Respondents on WASH, (n=396)

**Table 6:**
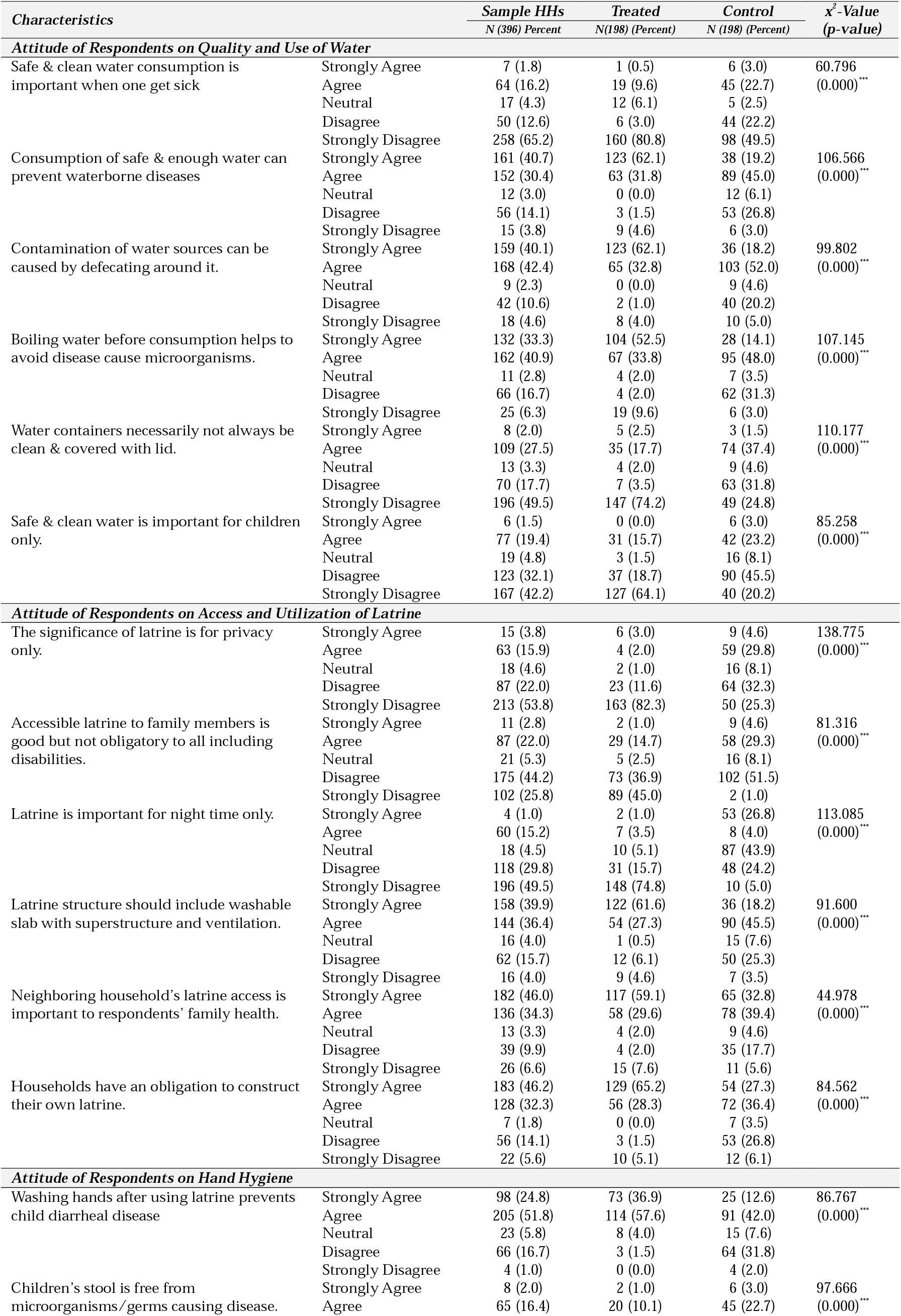
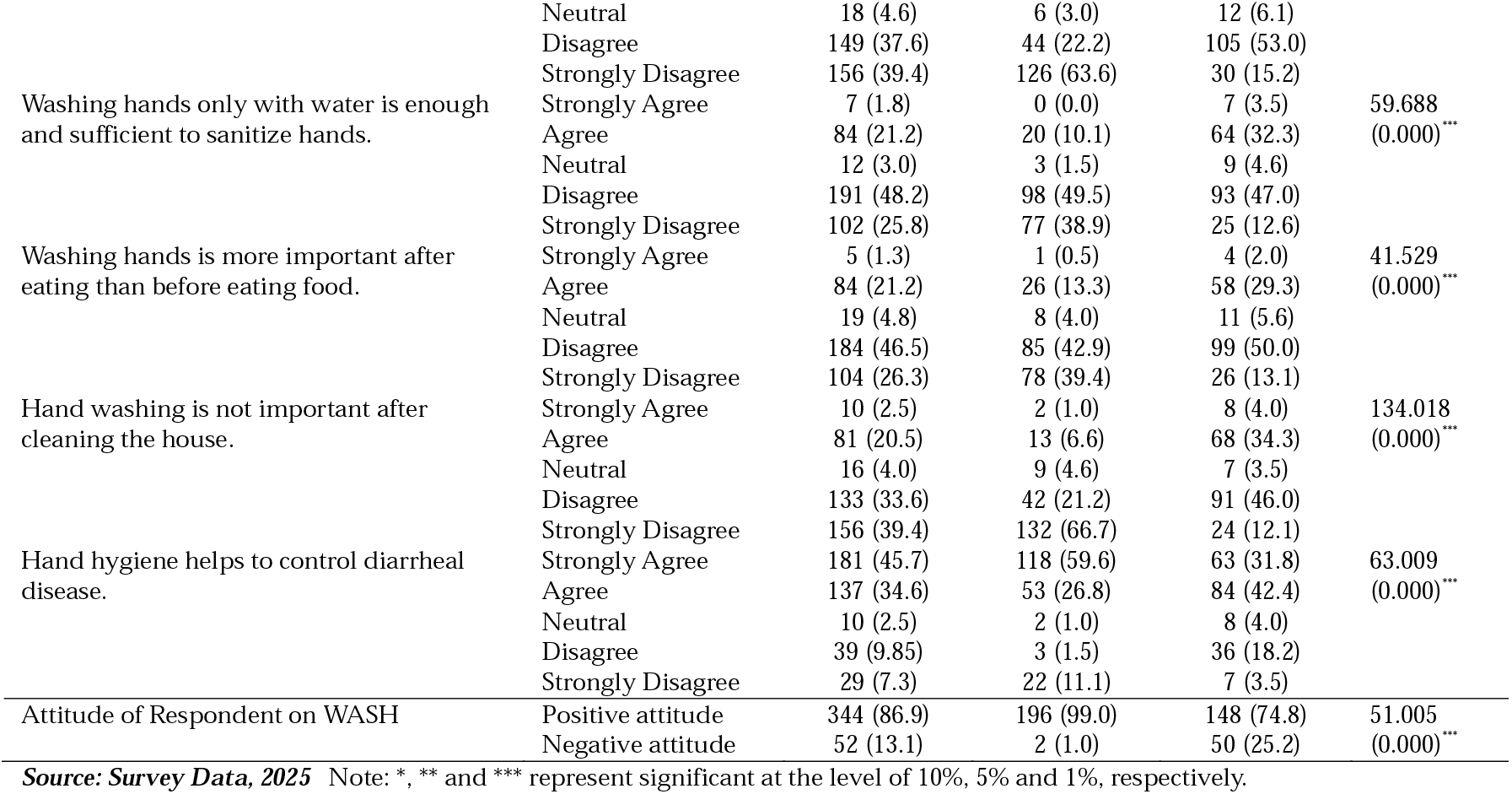
Attitude of Respondents on WASH.

**Table 7:** Practice of Respondents on WASH.

| Characteristics | | Sample HHs | Treated | Control | $\chi^2$-Value<br>(p-value) |
| --- | --- | --- | --- | --- | --- |
|  |  | N (396) Percent | N(198) (Percent) | N (198) (Percent) |  |
| Time taken to fetch water | ≤ 30 minutes | 265 (66.9) | 192 (97.0) | 73 (36.9) | 161.537 |
|  | > 30 minutes | 131 (33.1) | 6 (3.0) | 125 (63.1) | (0.000)*** |
| Solid waste disposal practice | Appropriate disposal | 256 (64.7) | 164 (82.8) | 92 (46.5) | 57.279 |
|  | Inappropriate disposal | 140 (35.3) | 34 (17.2) | 106 (53.5) | (0.000)*** |
| Household latrine utilization | Use properly all-time | 290 (73.2) | 177 (89.4) | 113 (57.1) | 52.766 |
|  | Not Use properly all-time | 106 (26.8) | 21 (10.6) | 85 (42.9) | (0.000)*** |
| Households have handwashing facility in the compound. | Yes | 227 (57.3) | 162 (81.8) | 65 (32.8) | 97.124 |
|  | No | 169 (42.7) | 36 (18.2) | 133 (67.2) | (0.000)*** |
| Materials used for handwashing | Water & soap or ash | 194 (49.0) | 145 (73.2) | 49 (24.8) | 93.129 |
|  | Water only | 202 (51.0) | 53 (26.8) | 149 (75.3) | (0.000)*** |
| Cleanliness of the household compound | Clean | 267 (67.4) | 143 (72.2) | 124 (62.6) | 4.150 |
|  | Unclean | 129 (32.6) | 55 (27.8) | 74 (37.4) | (0.042)** |
| Practice of WASH | Good | 221 (55.8) | 169 (85.4) | 52 (26.3) | 140.164 |
|  | Poor | 175 (44.2) | 29 (14.6) | 146 (73.7) | (0.000)*** |

**Source: Survey Data, 2025** Note: \*, \*\* and \*\*\* represent significant at the level of 10%, 5% and 1%, respectively.

### Attitude of Respondents on WASH

#### Attitude on Quality and Use of Water

As shown in Table 7, respondents in the treated group consistently showed more favorable attitudes toward safe water practices than the control group. Evidently, 83.8% of treated respondents valued clean water beyond illness, compared to 71.7% in the control group. Similarly, 93.9% of treated respondents supported drinking enough safe water to prevent waterborne diseases, versus 64.2% in the control group. Favorable attitudes were also higher in the treated group regarding contamination prevention near water sources (95.9% vs. 70.2%), boiling water before use (86.3% vs. 62.1%), keeping water containers clean and covered (77.7% vs. 56.6%), and recognizing that safe water is important for everyone (82.8% vs. 65.7%). Overall, the treated group demonstrated a stronger positive attitude across all WASH variables, reflecting the impact of the intervention.

#### Attitude on Access and Utilization of Latrine

Control group respondents showed higher unfavorable attitudes toward latrine importance and accessibility compared to the treated group. Clearly, 34.4% of control respondents viewed latrines as important only for privacy, versus 5% in the treated group. Similarly, 33.9% of control respondents disagreed that latrines should be obligatory for all family members, including those with disabilities, compared to 15.7% of treated respondents. Unfavorable attitudes toward nighttime latrine use were 30.8% in the control group and 4.5% in the treated group. Favorable attitudes on proper latrine structure were higher in the treated group (88.9%) than the control group (63.7%), and treated respondents were also more positive about neighbors’ latrine access (88.7% vs. 72.2%) and the obligation to construct household latrines (93.5% vs. 63.7%). Overall, the treated group exhibited stronger positive attitudes across all latrine-related indicators.

#### Attitude of Respondents on Hand Hygiene

Hand hygiene, critical for preventing waterborne diseases, showed much stronger positive attitudes in the treated group. About 94.5% of treated respondents washed hands after latrine use versus 54.6% in the control group. Favorable attitudes toward keeping children’s stools free of germs were 85.8% in the treated group and 68.2% in the control group. Across key handwashing misconceptions, such as washing with water alone, handwashing being more important after eating than before, and handwashing after cleaning the house, treated respondents consistently had higher positive responses (82–88%) compared to the control group (58–63%).

Overall, 99% of treated households had a favorable attitude toward water quality, latrine use, and hand hygiene, versus 74.8% in the control group. Only 1% of treated respondents held negative attitudes, compared to 25.2% in the control group, highlighting the impact of the project’s awareness campaigns and community-led sanitation initiatives.

#### Practice of Respondents on WASH

Table 7 shows significant differences in WASH practices between the groups. In the treated group, 97% fetched water within 30 minutes, compared to 36.9% in the control group, reflecting the impact of water infrastructure improvements. Proper solid waste disposal was reported by 82.8% of treated households versus 46.5% of control households. Latrine use was consistent in 89.4% of treated households, compared to 57.1% in the control group. Handwashing facilities were present in 81.8% of treated compounds versus 32.8% of control compounds, with 73.2% of treated respondents using soap or ash, compared to 24.8% in the control group. Compound cleanliness was higher in the treated group (72.2%) than in the control group (62.6%).

Overall, 85.4% of treated respondents had good WASH practices, while only 26.3% of control respondents did, with 73.7% of the control group showing poor practices, over three times higher than the treated group, highlighting the impact of hygiene campaigns and community-led sanitation interventions.

## Discussion

This cross-sectional study was conducted in Negelle-Arsi District of Oromia Regional State, Ethiopia, to assess water supply and access, sanitation and hygiene conditions, and the knowledge, attitudes, and practices (KAP) related to WASH among participants and non-participants of the WASH intervention. The WASH intervention was designed and implemented from 2018 to 2022 with the aim of reducing waterborne diseases, including child diarrhea. Diarrheal diseases are still one of the major causes of morbidity in children under five in sub-Saharan Africa [1]. It is among the top ten causes of death in Ethiopia in 2021, following lower respiratory infections and preterm birth complications, respectively. It is the cause of 33.67 deaths per 100 thousand population in total aged years. Access to at least a basic water supply service and improved sanitation contributes to human health and socio-economic development of a country [10].

The descriptive analysis findings show clear differences in WASH conditions and behaviors between households that participated in the intervention and those that did not. Important differences were also observed in child health– related practices. Mothers in the treated group were more likely to give birth in health facilities (88.9% vs. 63.6%), practice breastfeeding (98% vs. 89.9%), provide colostrum (99.5% vs. 87.9%), and vaccinate children against rotavirus (78.8% vs. 59.1%). These improvements correspond with a much lower prevalence of diarrhea among children in the treated group (8.1%) compared to the control group (37.4%).

Water access improved substantially in the intervention areas. All treated households used protected water sources compared to 76.8% of control households. The average time to fetch water was significantly shorter in the treated group (13.85 minutes) than in the control group (55.84 minutes), meeting the international standard of less than 30 minutes. Consequently, daily water consumption was almost double in the treated group (20.6 liters per person) compared to the control group (10.51 liters).This implies that improved water supply can reduce the time and effort required to fetch water. According to Jamie Cronk and Jamie Bartram (2018), such time savings lessen the burden on households, especially women and children, and can allow more time for education, income-generating activities, and other productive tasks [8]. Reliable and nearby water sources improve household well-being, reduce physical strain, and contribute to better health outcomes, including fewer cases of diarrhea and other hygiene-related diseases. These finding highlights that investing in local water infrastructure is an effective way to improve water access and support broader community health and development.

Sanitation conditions were also better in the treated communities. Latrine access was higher (95% vs. 78.3%), and open defecation was much lower (23.2% vs. 52%). Similarly, hygiene conditions improved, with 48.5% of treated households having handwashing facilities with soap and water compared to only 12.1% in the control group.

Knowledge, attitudes, and practices related to WASH were consistently stronger in the treated group. Good WASH knowledge was reported by 91.4% of treated households compared to about 70% of control households. Favorable attitudes toward WASH were observed in 99% of treated households and 74.8% of control households. In practice, 85.4% of treated households demonstrated good WASH practices compared to only 26.3% in the control group.

These differences highlight the effectiveness of awareness campaigns and community-led sanitation and hygiene approaches in improving knowledge, attitudes, and practices (KAP) related to WASH. Evidence also shows that combining infrastructure improvements with hygiene education and community mobilization significantly improves hygiene behaviors and reduces waterborne diseases [20]. Therefore, WASH interventions should integrate both infrastructure development and behavior change strategies to achieve sustainable health outcomes.

The results collectively demonstrate that the integrated WASH intervention significantly improved water access, sanitation, hygiene behavior, and child health outcomes in the intervention communities.

## Conclusions

Overall, the findings confirm that integrated WASH interventions, combining improved water infrastructure with sanitation promotion, hygiene education, and community engagement, can substantially improve household WASH conditions and reduce waterborne diseases. Sustaining these gains will require continued investment in water infrastructure, strengthening community awareness, and expanding behavior change initiatives to other communities.

## Data Availability

All data produced in the present study are available upon reasonable request to the authors

## Abbreviations

CLTSH: Community-led total sanitation and hygiene
JMP: Joint Monitoring Program
KAs: Kebele Administrations
KAP: Knowledge, Attitude & Practice
UNICEF: United Nation International Children’s Emergency Fund
WASH: Water, Sanitation and Hygiene
WHO: World Health Organization

## Acknowledgement

The authors acknowledge Negelle-Arsi District Water and Energy Office for their permission and support in collecting the data. Additionally, the authors acknowledge KOICA for funding the project for implementation and Habitat for Humanity Ethiopia for successfully implementing it.

## Authors Contributions

WF and BY: Both in conceptualization, study design, execution, acquisition of the data, analysis and interpretation, writing, and review and editing.

## Availability of data and materials

### Declarations

### Consent for publication

